# *De novo MAP2K4* variants cause a novel neurodevelopmental syndrome with impaired JNK signaling in iPSC-derived neurons

**DOI:** 10.64898/2025.12.23.25342932

**Authors:** Tomoki T. Nomakuchi, Alyssa L. Rippert, Sabrina A. Santos De León, Elizabeth M. Gonzalez, Dong Li, Rajesh Angireddy, Livia Sertori Finoti, Flavio Faletra, Luciana Musante, Rinne Tuula, David J. Amor, Lydia von Wintzingerode, Rami Abou Jamra, Samantha R. Stover, Jillian G. Buchan, Jennifer Hayek, Eyby Leon, Tania Attie-Bitach, Marlene Rio, Genevieve Baujat, Elisabeth Wallach, Amandine Smail, Kerith-Rae Dias, Ulrich Pfeifer, Amanda Peterson, Rebecca C. Ahrens-Nicklas, Elizabeth J. K. Bhoj

**Author notes:** Authors contributed equally (Tomoki T. Nomakuchi, Alyssa L. Rippert, Rebecca C. Ahrens-Nicklas, Elizabeth J. K. Bhoj).

## Abstract

*MAP2K4* encodes a kinase that activates the c-Jun N-terminal kinase (JNK) pathway, which is essential for human neurodevelopment. While somatic MAP2K4 loss has been observed in cancer, germline variants have not previously been linked to human disease. We describe ten individuals with *de novo* or presumed *de novo MAP2K4* variants who present with a novel syndromic neurodevelopmental disorder. Shared features include developmental delay or intellectual disability, epilepsy, and variable congenital malformations, most commonly affecting the genitourinary system. To define the mechanism, we generated CRISPR-edited iPSC-derived neurons with MAP2K4 deficiency. These neurons showed reduced JNK pathway activation and abnormal differentiation, characterized by persistence of progenitor-like cells and disrupted neurite morphology. Our findings establish *MAP2K4* as a Mendelian neurodevelopmental disorder gene and identify impaired JNK signaling as the underlying mechanism. More broadly, this work expands the spectrum of JNK-pathway disorders and underscores the critical role of JNK signaling in human brain development.

## Introduction

MAP2K4 is a dual-specificity kinase that canonically activates the c-Jun N-terminal kinases (JNK) pathway in response to a variety of cellular stimuli, including mitogenic or cell death signals, oxidative stress, and pro-inflammatory signals^1,2^. MAP2K4, along with MAP2K7, phosphorylates JNK1, JNK2, and JNK3. MAP2K4 can also phosphorylate and activate the p38 kinase in a context-dependent manner, along with MAP2K3, MAP2K6, and additional noncanonical pathways^3,4^. The known role of MAP2K4 in human diseases has been largely limited to the context of cancers, where recurrent loss of MAP2K4 is observed and has been proposed as possible driver in several malignancies^5,6^. *MAP2K4* is highly expressed in the human brain at all developmental stages, and evolutionary constraint metrics suggest that it is loss-intolerant with pLI of 1.0, LOEUF of 0.22 and pHaplo of 0.97^7–9^. Knockout of *Map2K4* in mice was shown to result in early embryonic lethality, and conditional knockout in the central nervous system resulted in severe brain malformations^10,11^. In a recent genome-wide prediction study of neurodevelopmental disorder risk genes, *MAP2K4* was highlighted as a candidate gene for monoallelic NDD phenotypes^12^.

Several key regulators of the JNK pathways are linked with human Mendelian disorders. *De novo* truncating and missense variants within *MAPK8IP3* have been reported in individuals with severe syndromic neurodevelopmental disability (NDD) with brain malformation^13^. This gene encodes the JNK-interacting protein 3 (JIP3), which is involved in the axonal transport of cargos including lysosomes and activated JNK^14,15^. Likewise, germline variants within the *MAP4K4* gene were recently linked with NDD and multiple congenital anomaly syndrome^16^. The MAP4K4 protein is an upstream regulator of multiple kinase pathways including the JNK pathway^17^.

Data from cell and animal models, as well as emergence of novel Mendelian syndromes linked with the JNK pathway, suggest that appropriate regulation of the JNK pathway is necessary for normal human neurodevelopment^18–20^. Despite its key role in the direct phosphorylation and activation of JNK, germline variants in *MAP2K4* have not been previously associated with a human disease phenotype. Here, we present 10 individuals with syndromic NDD and germline variants in *MAP2K4*, most of which were confirmed *de novo*. We additionally modeled heterozygous and biallelic loss of MAP2K4 in iPSC-derived neurons, demonstrating reduction in the activation of the JNK pathway as well as abnormal neuronal differentiation. Together, these clinical and functional findings establish *MAP2K4* as a novel Mendelian NDD gene, expanding the spectrum of JNK-pathway–related syndromes.

## Methods

### Patient ascertainment and clinical data

Probands were referred from clinical genetics centers following exome or genome sequencing. Exome or genome sequencing was performed using a variety of standard clinical capture kits and platforms.

Referral and assembly of the cohort was facilitated by GeneMatcher^21^. The prenatal, perinatal, developmental, neurologic, and systemic findings, as well as photographs when available, were reviewed for inclusion in the cohort by three clinical geneticists (TTN, RAN, EB) and two molecular geneticists (DL and EB). Written informed consent was obtained under institutional review board–approved protocols.

### Genetic testing and variant interpretation

Exome or genome sequencing was performed on proband–parent trios (n=9) or proband-mother duo (n=1) using various standard exome or genome platforms and standard clinical and research pipelines. Variants were annotated and interpreted according to American College of Medical Genetics and Genomics/Association for Molecular Pathology (ACMG/AMP) guidelines.

### Generation of MAP2K4-deficient human iPSCs

iPSCs were maintained as feeder-free cultures with mTeSR Plus Medium (StemCell Technologies, 100-0274) on hESC-qualified Matrigel (Corning, 356234), with media change every 1-2 days. MAP2K4-deficient iPSC cells were generated from parental NIH CRM control iPSC line (NCRM-1)^22^ through CRISPR/Cas9-mediated introduction of random insertions/deletions within exon 7. Guide RNA sequence and primer sequence are included within Supplemental Table 1. The gRNA was cloned into Cas9/gRNA dual-expression vector (Addgene, 134451)^23^ and transfected using Lipofectamine Stem Transfection Reagent (Thermo Fisher STEM00001) according to the manufacturer protocol. Effectively transfected cells were selected using puromycin at 0.5 µg/mL for 48 hours. Single clones were selected and expanded.

Sanger sequencing (CHOP Center for Applied Genomics) and next-generation sequencing (Plasmidsaurus) were performed at the target site within exon 7 to assess for edits and to confirm monoclonal population. SNP microarray was performed using the Infinium Global Screening Array v3.0 Kit (Illumina) on all edited clones. Microarray data was analyzed on the GenomeStudio software (Illumina). The OMIM-annotated disease genes affected by copy number variants were identified using GeneScout (https://genescout.omim.org/).

### Protein extraction and Western blot

Cells were lysed and protein isolation was performed in RIPA buffer containing protease and phosphatase inhibitor cocktail (Thermo Fisher, 78440), and protein concentration in the lysate was measured using the BCA Protein Assay Kit (Thermo Fisher, 23225). 15μg of protein was loaded onto the NuPAGE 4–12% Bis-Tris Protein Gel (Thermo Fisher, NP0322) and separated via electrophoresis. The proteins were transferred onto PVDF membranes using the iBlot™ 3 Western Blot Transfer Device (Thermo Fisher) using the “Broad Range” setting. The membranes were blocked with 5% bovine serum albumin for 1hr at room temperature. The membranes were incubated overnight at 4 °C with the following primary antibodies and dilutions: rabbit anti-MAP2K4 (Cell Signaling, 9152, 1:1000), rabbit anti-JNK (Cell Signaling Technology, 9252, 1:1000), mouse anti-phosphorylated JNK (Cell Signaling Technology, 9255, 1:500), rabbit anti-vinculin (Abcam, ab91459, 1:2000). Membranes were washed three times in TBST followed by incubation in secondary antibodies at 1:10,000 (IRDye 680RD goat anti-rabbit (LI-COR, 926-68071) and IRDye 800CW goat anti-mouse (LI-COR, 926-32210)). The blots were visualized using the LICOR Odyssey system and the band intensities were analyzed using ImageJ.

### NGN2-mediated differentiation of iPSCs into neurons

NGN2-induced neurons (iN) were generated as previously described^24^. Briefly, NGN2-lentivirus was generated in HEK293T cells by co-transfection of lentiviral packaging and envelope plasmids pMDLg/pRRE (Addgene,12251), pRSV-Rev (Addgene,12253), and VSV-G (Addgene, 12259), along with NGN2 expression plasmid pLVX-UbC-rtTA-Ngn2:2A:EGFP (Addgene, 127288). Conditioned media was collected 48 hours post transfection and filtered through a 0.45 μm filter.

iPSCs were transduced with the lentiviral constructs in the presence of 10 μM ROCK inhibitor Y-27632 (Tocris, 1254). 24 hours following transduction, transduced cells were selected with puromycin at 0.5 µg/mL, followed by single clone selection. Following expansion of individual clones, successful and durable transduction with the NGN2 construct was confirmed with a second round of puromycin selection as well as expression of GFP upon addition of doxycycline.

NGN2-iPSCs were dissociated with accutase (Sigma-Aldrich, A6964) and seeded at 700,000 cells per well on Matrigel-coated 6-well plates in pre-differentiation Medium (Knockout DMEM/F12 (Gibco, 12660–012), N2 supplement (Gibco, A1370701), MEM Non-Essential Amino Acids (Gibco, 11140–050)) supplemented with 10 μM ROCK inhibitor, 0.2 μg/mL Mouse Laminin (Thermo Fisher, 23017–015), 10 ng/mL NT-3 (PreproTech, 450–03), 10 ng/mL BDNF (PreproTech, 450–02), and 2 μg/mL doxycycline (Sigma-Aldrich, D3072). Media was replenished daily. After 3 days, the pre-differentiated cells were transferred to plates coated with poly-L-ornithine (Sigma-Aldrich, P3655) and Matrigel at a density of 200,000 cells per 6-wells in Classic Neuronal Medium (Neurobasal Plus Medium (Gibco, A3582901), N2, B27 (Gibco, 17504–044), MEM Non-Essential Amino Acids, Glutamax (Gibco, 35050–061)) supplemented with 2 μg/mL doxycycline hydrochloride, 10 ng/mL NT-3, 10 ng/mL BDNF and 1 μg/mL mouse laminin. Cells for immunocytochemistry were plated onto poly-D-lysine coated coverslip (Corning, 354087) additionally coated with Matrigel. Partial media change without doxycycline was performed every 4-5 days. The cells were prepared for protein extraction, RNA extraction or immunostaining at day 11 of differentiation.

### Immunofluorescence

Cells were fixed with 4% paraformaldehyde for 15min at room temperature, then washed gently three times with PBS. Cells were then permeabilized with 0.1% Triton X-100 (Sigma-Aldrich, T8787) in PBS and blocked for 2 hours at room temperature with 5% Normal Goat Serum (Jackson Immuno, 005–000-121) in PBS. Slides were then incubated overnight at 4 °C in primary antibodies. The following antibodies and dilutions were used: mouse anti-Tubulin β3 (R&D Systems, 1195) at 1:800, rabbit anti-MAP2 (Cell Signaling Technology, 4542) at 1:500, mouse anti-phosphorylated JNK (Cell Signaling Technology, 9255) at 1:200, rabbit anti-JNK (Cell Signaling Technology, 9252) at 1:200. The slides were then washed 3 times in PBS and incubated in secondary antibodies (Alexa Fluor Plus 555 Donkey anti-rabbit (Thermo Fisher, A32794) and Alexa Fluor 647 donkey anti mouse (Thermo Fisher, A31571)) at 1:1000 dilution for 1 hour. Slides were then stained with DAPI (NucBlue Fixed Cell Stain ReadyProbes reagent (Thermo Fisher, R37606) at 1 drop per 500ul) for 10 minutes, followed by two washes in PBS prior to mounting. Images were acquired using a Keyence BZ-X800. Fluorescence image intensities were quantified using a custom Python script. Raw TIFF files from each channel were read with tifffile, and the mean pixel intensity was calculated for each channel across the full image field.

### RNA isolation and qPCR

Total RNA was isolated from cells using Direct-zol RNA Miniprep kit (Zymo, R2050) according to manufacturer’s instructions. cDNA was prepared using the High-Capacity RNA-to-cDNA kit (Applied Biosystems, 4387406). qPCR was performed using PowerUP SYBR Green Master Mix (Thermo Fisher, A46012) on a QuantStudio 3 Real-Time PCR System (Applied Biosystems), using *GAPDH* as the control housekeeping gene. Expression relative to the control cells were analyzed using the ΔΔCt method.

### Statistical analysis

All experiments were performed with at least three independent biological replicates. Statistical analyses were performed using GraphPad Prism. Analysis between the CRISPR-edited clones were done using one-way ANOVA with Fisher’s LSD post-hoc testing. Statistical significance was indicated as follows: *p < 0.05, **p < 0.01, ***p < 0.001, ****p < 0.0001

## Results

### Cohort demographics and variant spectrum

We have identified ten probands (six male, four female; median age at last assessment 10 years). Five variants were predicted loss-of-function (c.495_513+9del; c.733_737del, p.Glu92Ter, p.Leu245*; p.Arg281*), and five were missense or in-frame indels that affected residues within the kinase domain (Table 1 and Figure 1). Nine of ten variants were confirmed *de novo*. One proband underwent a duo exome including the proband and the mother, and the variant was not detected in the maternal DNA.

**Figure 1.**
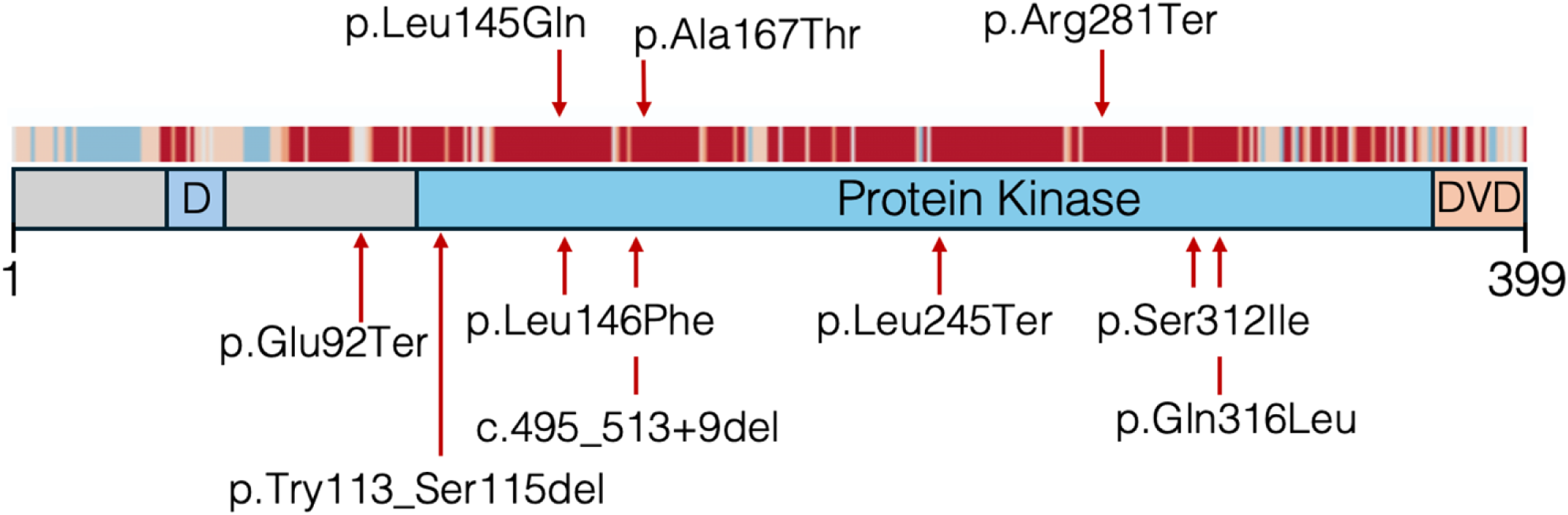
The types and locations of the variants identified in this study, positioned on the schematic of the MAP2K4 protein. AlphaMissense scores as heatmap obtained from UniProt is positioned above the protein diagram, with red color representing positions where missense variants are likely damaging. The D-domain, kinase domain and the DVD domain are annotated.

### Prenatal and perinatal findings

Eight of ten pregnancies (80%) had complications including polyhydramnios, oligohydramnios, intrauterine growth restriction, and increased nuchal translucency. One pregnancy was terminated due to multiple anomalies. Two infants suffered perinatal stroke; one of these infants died in the neonatal period secondary to perinatal stroke and prematurity. Neonatal issues included hypoglycemia, jaundice, and umbilical infection.

### Neurodevelopmental and neuroimaging findings

Developmental delay or intellectual disability (ID) was documented in 7 of 8 evaluated children (87.5%). Median age at sitting was 9.25 months, walking at 20 months, and first words at 21.4 months. Four probands had formal IQ testing with median IQ of 63 (range 55-66). Behavioral issues (such as self-injurious behaviors, sensory issues, frustration intolerance, or aggressivity) were noted in 3/8 (37.5%) children, and one was formally diagnosed with autism. Brain MRI (n=8) revealed focal cortical dysplasia type II (n=1), corpus callosum abnormalities (n=2), nonspecific white matter changes (n=1), intraventricular hemorrhage (n=1), ventriculomegaly (n=1), and cerebellar atrophy (n=1). Four of eight carried a diagnosis of epilepsy (50%).

### Additional organ involvement

Musculoskeletal anomalies were particularly common (6/8, 75%) and included scoliosis, hip dysplasia, camptodactyly, talipes, vertebral anomalies, delayed bone age, and short stature (3/8, 37.5%).

Genitourinary anomalies were present in 5/10 (50%), including ectopic kidney, pyelectasis, hypospadias, and megacystis. Seven of nine evaluated for dysmorphology showed variable dysmorphic features such as clinodactyly, down-slanting palpebral fissures, long philtrum, and webbed neck (Fig. 1B-C).

### Generation of MAP2K4-deficient iPSCs

Frame-shifting indels were introduced in the coding region of exon 7 of *MAP2K4* using CRISPR/Cas9, in the NCRM1 reference iPSC^22^. The exon 7 contains a portion of the kinase domain, and the patient-specific p.Leu245Ter variant is located within this exon (Figure 1A). Clones of edited iPSCs were isolated and sequenced to identify clones with heterozygous and biallelic frameshift variants. The control line was transfected with CRISPR/Cas9 without gRNA and underwent the same clonal selection process (Clone ‘Cas9’). Two clones with heterozygous edits as well as one clone with biallelic edits were selected for further experiments (Clones ‘Het 1’, ‘Het 2’ and ‘KO’; Fig. 2A). Western blot confirmed reduction of MAP2K4 protein levels in the heterozygous clones, and absence in the biallelic clone (Fig. 2B). These clones demonstrated reduced *MAP2K4* transcript levels, presumably from nonsense-mediated mRNA decay (Fig. 2C). Genomic DNA from each of the clones were sent for chromosomal microarray, which demonstrated multiple microdeletions and duplications largely shared between all clones including the Cas9-control clone. Copy number variants (CNVs) greater than 10kb are reported in Supplementary Table 1 along with disease-associated genes withing the regions. Deletions primarily involved tumor-associated genes including *APC*, *ATM*, *MSH2*, and *RB1* shared across all four clones, with others including *STK11* and *BRCA2* present in some but not all clones. NDD and epileptic encephalopathy genes were among the deleted genes, including *SCN1A* (Dravet syndrome, OMIM# 607208) deleted in all clones and *NIPBL* (Cornelia de Lange syndrome, OMIM# 122470) deleted in the Cas9, Het 1 and Het 2 clones but not in the KO clone.

**Figure 2.**
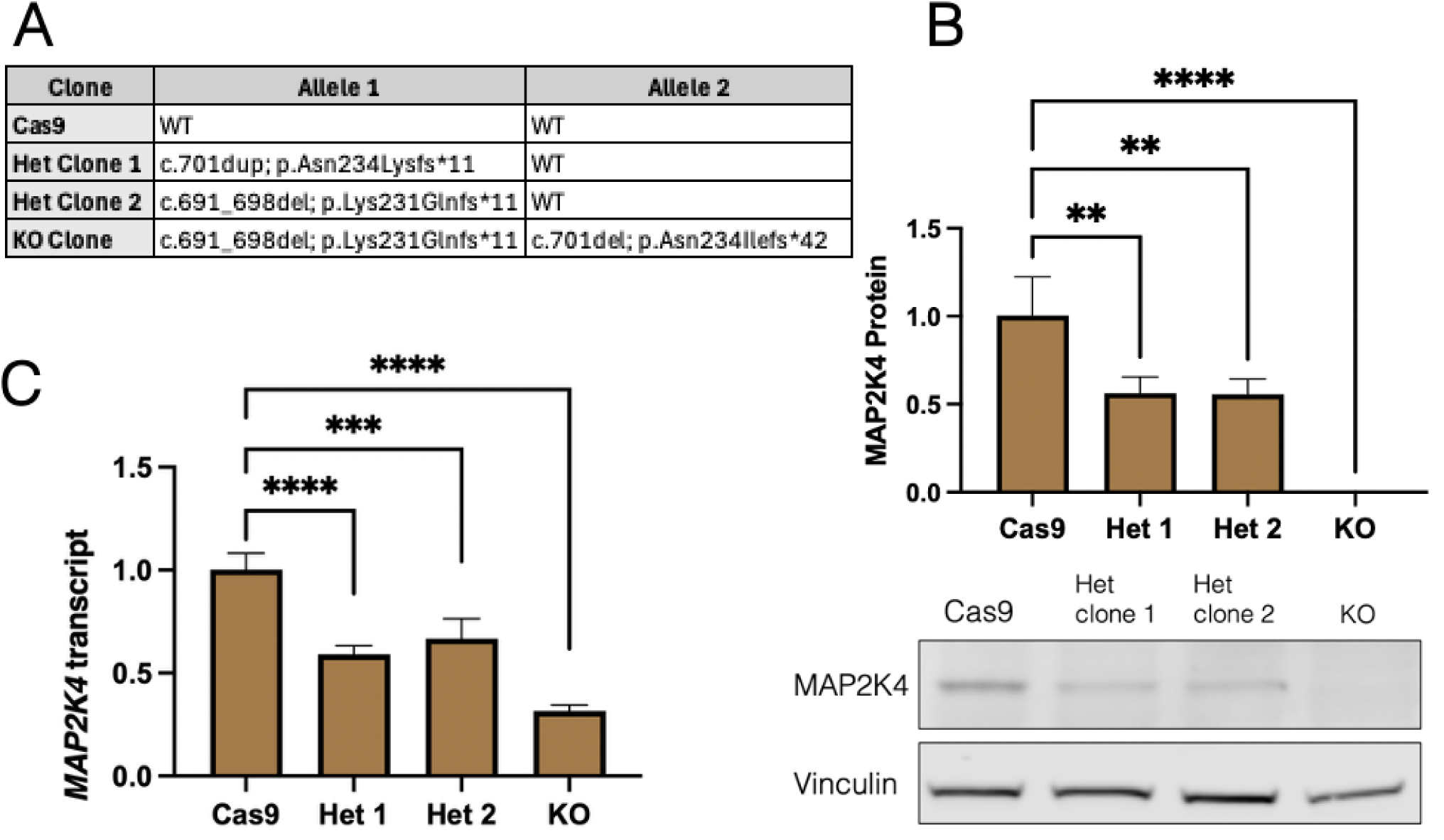
A) Mutations introduced in the NCRM1-iPSCs used in our study. The cells with frameshift mutations in MAP2K4 demonstrated expected reduction in both B) protein level and C) mRNA level (n=3).

### Characterization of iPSC-derived neurons

Given the high prevalence of NDD, epilepsy and abnormal brain MRI findings, we next modeled MAP2K4 deficiency in iPSC-derived neurons. We used a lentiviral neurogenin-2 (NGN2) overexpression system to rapidly generate NGN2-induced neurons (NGN2-iN)^24,25^. Each clone demonstrated robust differentiation into cells expressing both beta-tubulin III and MAP2 after 11 days of differentiation (Fig. 3). The MAP2K4-deficient cells, however, demonstrated disorganized axonal architecture compared to the Cas9 control, and more dense clusters of nuclei, most evident in the KO clone (Fig. 3, bottom). Ki-67 staining demonstrated that much of clustered cells are Ki-67 positive, indicative of a proliferative state (Fig. 4A-B). The expression of neural progenitor marker *PAX6* was markedly increased in the edited clones, indicating that at least some of the proliferating cells are neural progenitor-like cells (Fig. 4C). Conversely, expression of *MAP2* was overall reduced, consistent with a mixed cell population rather than pure neuronal culture (Fig. 4C).

**Figure 3.**
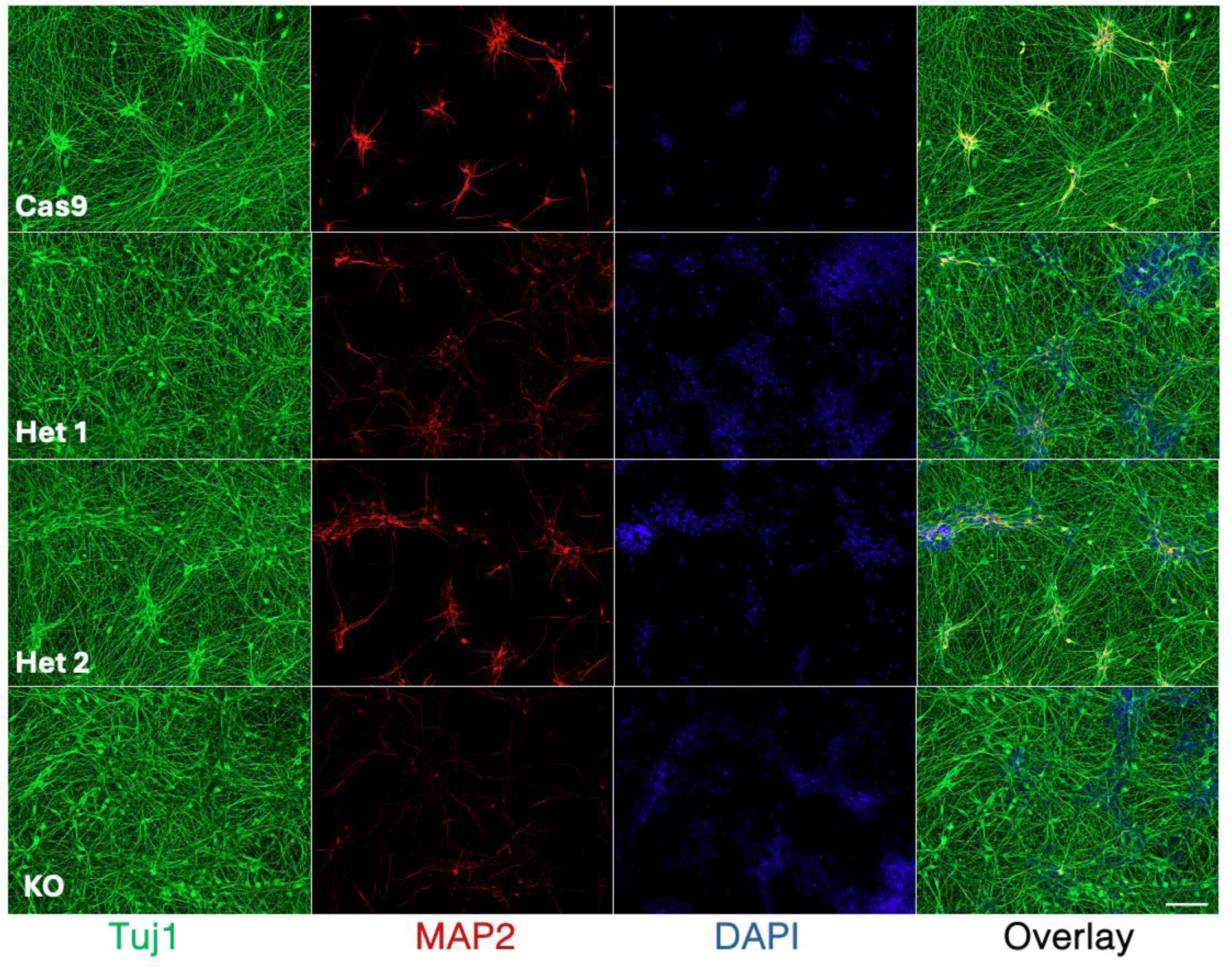
Differentiation of iPSC-derived iNs were characterized with immunofluorescence for Tuj1 (Beta-III tubulin) and MAP2. Scale bar = 100µm. Note the increased cellularity in the Het 1, Het 2 and KO clones, as well as disorganized axonal architecture in the KO clone.

**Figure 4.**
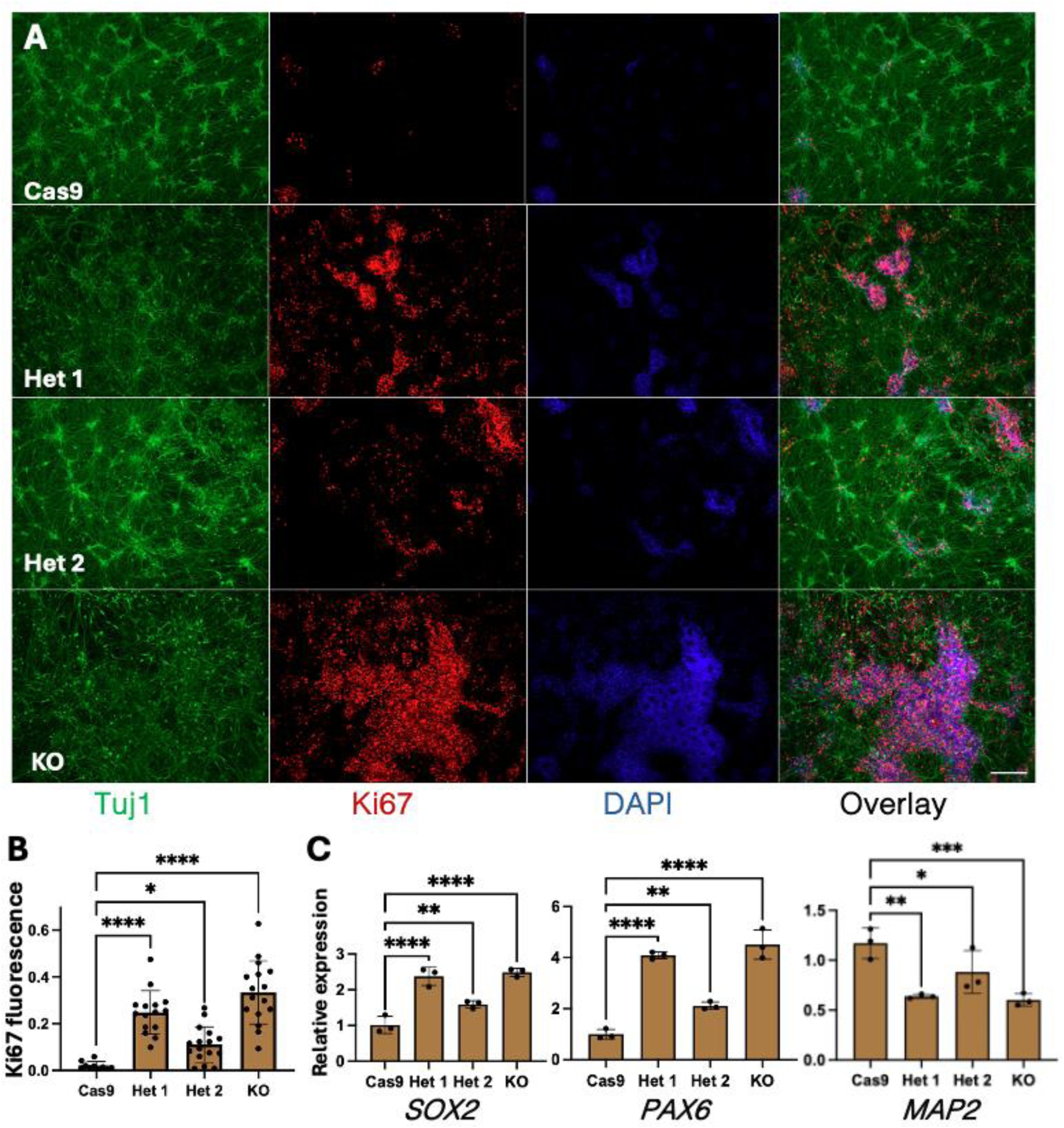
**A)** Immunofluorescence of iNs stained for Tuj1 and Ki67, demonstrating abundance of proliferating cells in the MAP2K4 deficient cells. Scale bar = 200µm. B) Quantification was done with mean fluorescence of Ki67 normalized for Tuj1 fluorescence (n=10). C) qPCR of iNs at day 11 of differentiation demonstrated elevated expression of NPC markers *SOX2* and *PAX6*, whereas mature neuronal marker *MAP2* is reduced in MAP2K4 deficient cells.

We next examined the activation of the JNK pathway in the MAP2K4-deficient neurons. Co-staining of total JNK and phosphorylated JNK demonstrated reduced JNK phosphorylation in all three edited clones, although residual phosphorylation of JNK was observed in all clones including the biallelic KO clone (Fig. 5A-B). JNK activates the transcription factor c-Jun, which can upregulate its own transcription among other transcriptional targets^26^. The iNs demonstrated reduced *JUN* expression, consistent with reduced JNK activation (Fig. 5C).

**Figure 5.**
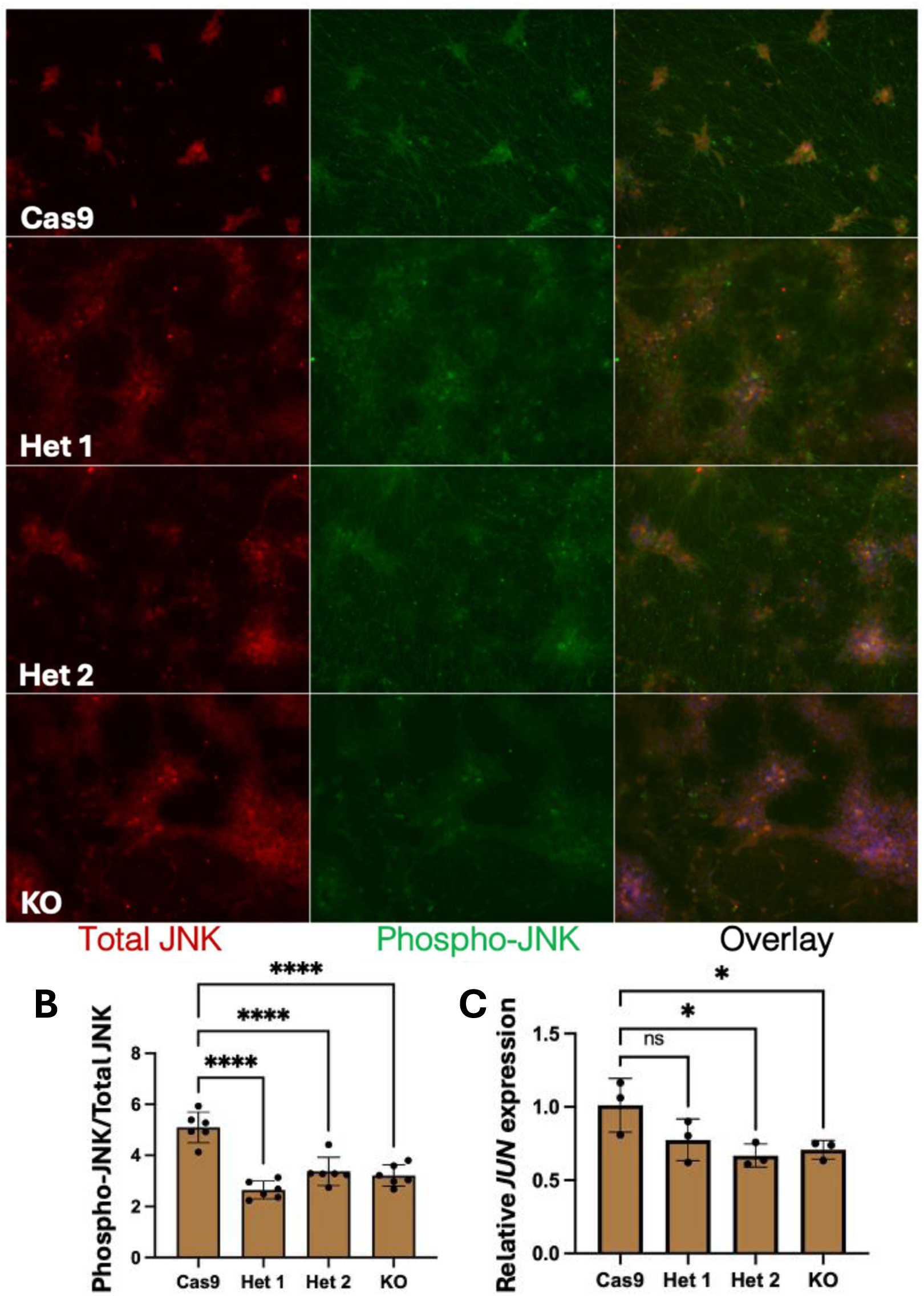
**A)** Immunofluorescence of total (red) and phosphorylated (green) JNK, quantified as mean fluorescence of phospho-JNK normalized for total JNK fluorescence, demonstrating reduction of phosphor-JNK in MAP2K4 deficient iNs (n=5), quantified in (B). Scale bar = 200µm. C) qPCR of iNs at day 11 of differentiation demonstrated reduction in *JUN* expression in MAP2K4 deficient cells (n=3).

## Discussion

Here, we report a novel syndromic NDD caused by *de novo MAP2K4* variants. The clinical features in our cohort included ID in the majority of living probands (7/8), structural brain anomalies on MRI in a subset, and epilepsy in half. Congenital malformations were frequent, particularly musculoskeletal (scoliosis, hip dysplasia, camptodactyly, vertebral anomalies) and genitourinary anomalies (ectopic kidney, megacystis, pyelectasis, hypospadias). Cardiac anomalies were less common and included ventricular septal defect and patent ductus arteriosus. Prenatal complications such as polyhydramnios, oligohydramnios and fetal growth restriction were observed in most pregnancies, and perinatal stroke occurred in two probands. Dysmorphic features were variable and nonspecific but often included down-slanting palpebral fissures, long philtrum, and coarse facies. Our functional data confirms the reduction of JNK activation in cell lines with *MAP2K4* haploinsufficiency, and abnormal neuronal differentiation.

In addition to highlighting *MAP2K4* as a novel Mendelian neurodevelopmental disorder gene, our findings expand the spectrum of neurodevelopmental disorders linked to the JNK pathway. Mendelian NDD syndromes due to genes linked with the JNK pathway include *MAP4K4*, *MAPK8IP3*, *TAOK1* and *TAOK2*-related NDD^13,16,27^. These syndromes share NDD phenotype as their core feature, with or without additional findings including structural brain anomalies and epilepsy. Because they have only recently been described, the full spectrum and defining characteristics of their NDD phenotypes, as well as their underlying mechanisms including the role of the JNK pathway, remain to be determined. MAP4K4, for instance, can regulate JAK-STAT, Notch, NF-8kB and MAPK/ERK pathways^16,17^. TAOK1 can likewise modulate Hippo/YAP/TAZ signaling in addition to p38 and JNK pathway activation, and TAOK2 appear to regulate primary cilia length^28,29^. Mechanistic data are needed to define the contribution of altered JNK pathway in these disorders.

Our edited iNs demonstrated impaired phosphorylation of JNK and reduced expression of *JUN*, a key autoregulatory target of activated c-Jun. This impaired activation coincided with defective neuronal differentiation in vitro, characterized by persistence of proliferative progenitor-like cells, reduced neuronal marker expression, and, most evident in the KO clone, disorganized neurite morphology.

Residual JNK phosphorylation in the biallelic knockout line suggests partial pathway redundancy, likely mediated through MAP2K7 or other kinases. Importantly, the JNK pathway is activated under a variety of cellular stressors, including inflammation, hypoxia, hyperthermia and glucocorticoid stimulation, all of which are highly relevant in the context of neurodevelopment^30–33^. Further work is needed to determine whether extracellular stressors modify the observed phenotype in *MAP2K4*-edited cells or possibly uncover additional, stress-dependent phenotypes. Such findings could relate back to the variable phenotype observed in the patients and suggest approaches to minimize the disease burden.

Our study has several limitations, including the deletions of key neurological disease genes observed in the iPSC clones. This limitation was partially mitigated by the use of a control clone with the same CNVs. CNVs associated with NDDs have been reported in an independent iPSC line^34^, and may be an underrecognized and common consequence of iPSC maintenance in culture. Additional limitations of our study include modest patient cohort size and limited follow-up time, and limited information for two probands in whom pregnancy ended in termination or intrauterine demise. Functional studies were limited to the NGN2-iN model, which is an appropriate model for early excitatory cortical neurons, but do not capture the full diversity of cell types affected *in vivo*.

Taken together, our findings define MAP2K4 as a novel etiology of syndromic NDD and nominate the JNK pathway as the molecular etiology to this phenotype. Expansion and long-term observation of affected individuals will be essential to refine the clinical spectrum and assess genotype–phenotype correlations. Based on the recurrent findings in this cohort, we recommend that newly diagnosed patients with MAP2K4-related disorder undergo a focused baseline evaluation. Given the occurrence of congenital heart disease, we suggest an echocardiogram at diagnosis. Because renal anomalies were present in half of the probands, renal ultrasound should be obtained. Musculoskeletal anomalies are common, but typically apparent on exam. Neurodevelopmental evaluation with referral for early intervention and developmental specialists should be considered in all patients. The high risk of seizures should be discussed at the time of diagnosis, and a low threshold for formal neurological evaluation is recommended. Brain MRI may be considered if there are seizures or neurologic concerns. Although loss of *MAP2K4* is recurrently observed in tumors^5^, history of malignancies was absent in our cohort. Long-term follow-up and cohort expansion will be important to determine whether additional surveillance could be warranted, including screening for malignancies.

Additional mechanistic studies with alternate models such as cerebral organoids, independent iPSC lines, and animal models are needed to better define how decrease of MAP2K4 and perturbation of the JNK pathway underlie NDD pathogenesis. Interestingly, activation of the JNK pathway has also been implicated in neurodegenerative diseases including Parkinson’s and Alzheimer’s, where JNK inhibition is under investigation as a therapeutic approach^35^. Our description of MAP2K4-related NDD further underscores the context-dependent but critical role the JNK pathway plays in early neurodevelopment as well as later-onset neurodegeneration.

## Data Availability

All data produced in the present study are available upon reasonable request to the authors

## Acknowledgments

T.T.N. was supported by NIH K08 (NS140553) and NIH T32 (GM008638). This work was funded in part by a grant from the Chan Zuckerberg Initiative to E.B. and R.C.A.-N.

---

| Proband | 1 | 2 | 3 | 4 | 5 | 6 | 7 | 8 | 9 | 10 |  |
| --- | --- | --- | --- | --- | --- | --- | --- | --- | --- | --- | --- |
| Variant detail (NM_003010.4) | c.733_737del | c.935G>T | 436C>T | c.947A>T | c.841C>T | c.495_513+9del | c.337_345del | c.274G>T | c.434_435delinsAA | c.425A>C |  |
|  | p.Leu245_Cys246delins* | p.Ser312Ile | p.Leu146Phe | p.Gln316Leu | p.Arg281* | p.? | p.Try113_Ser115del | p.Glu92* | p.Leu145Gln | p.Gln142Pro |  |
| Inheritance | not maternally inherited | de novo | de novo | de novo | de novo | de novo | de novo | de novo | de novo | de novo |  |
| Age range (years) | 5-10 | 5-10 | Pregnancy terminated | 10-15 | 10-15 | Intrauterine demise | 5-10 | 10-15 | 15-20 | 10-15 |  |
| Sex | Male | Male | Male | Female | Male | Male | Female | Male | Female | Female |  |
| Developmental History |  |  |  |  |  |  |  |  |  |  | Overall Frequency |
| Developmental Delay | Yes | No | N/A | Yes | Yes | N/A | Yes | Yes | Yes | Yes | 7/8 (87.5%) |
| Intellectual Disability | N/A | No | N/A | Yes | Yes | N/A | Yes | Yes | Yes | Yes | 6/7 (85.7%) |
| Autism | Yes | No | N/A | No | No | N/A | No | No | No | No | 1/8 (12.5%) |
| ADHD | Yes | No | N/A | No | No | N/A | No | No | Yes | No | 2/8 (25%) |
| Behavior Concerns | Yes | No | N/A | No | No | N/A | No | Yes | Yes | No | 3/8 (37.5%) |
| Neurologic Findings |  |  |  |  |  |  |  |  |  |  |  |
| Hypotonia | Yes | No | N/A | No | No | N/A | Yes | No | Yes | No | 3/8 (37.5%) |
| Epilepsy | No | No | N/A | Complex partial seizures | Focal impaired awareness, motor onset | N/A | Lennox-Gastaut syndrome | No | Generalized epilepsy | No | 4/8 (50%) |
| EEG Findings | Normal | Normal | N/A | N/A | sporadic sharp waves | N/A | Multifocal discharges with background slowing | N/A | N/A | No | 2/5 (40%) |
| Brain MRI Abnormalities | Focal cortical dysplasia, undulating contour to the left anterior body of corpus callosum | N/A | N/A | No | Nonspecific white matter changes | Multiple and expansive germinal matrix hemorrhage | Occipital encephalomalacia, ventriculomegaly | No | Thin corpus callosum, cerebellum moderate atrophy (vermis and lobes), abnormal temporal lobes | No | 5/8 (62.5%) |
| Organ System Involvement |  |  |  |  |  |  |  |  |  |  |  |
| Ophthalmologic | Astigmatism | Hypermetropia | N/A | N/A | N/A | N/A | Exotropia | Astigmatism | Astigmatism | Amblyopia | 6/6 (100%) |
| ENT/Hearing loss | Recurrent ear infections | delayed tooth eruption | N/A | Mild sensorineural hearing loss | No | N/A | No | No | N/A | Delayed tooth eruption | 4/7 (57.1%) |
| Respiratory | Obstructive sleep apnea necessitating tonsillectomy/adenoidec tomy | No | N/A | Yes | N/A | N/A | Restrictive lung disease, obstructive apnea | No | No | No | 3/7 (42.3%) |
| Cardiac | No | No | Persistent superior vena cava | Pulmonary artery stenosis, ASD, PFO | No | Pleural & pericardiac effusion | No | No |  | No | 3/9 (33.3%) |
| Gastrointestinal | Selective diet and swallowing issues | No | Absent gallbladder | No | N/A | N/A | Gastrostomy tube dependent | No | Yes | Selective diet | 4/7 (57.1%) |
| Genitourinary | Ectopic kidney | right pyelectasis with dilated calyces & cortical thinning | Hypospadias | No | No | Megacystis w/o obstruction, hydronephrosis | No | Mild intermittent hydronephrosis | No | No | 5/10 (50%) |
| Musculoskeletal | No | Short stature with delayed bone age | Clenched fists | Talipes, narrow chest, severe scoliosis | Pes equinus | N/A | Hip dysplasia, scoliosis | Short stature, scoliosis, butterfly vertebra, contracures of fingers and toes s/p surgical correction | Joint stiffness, camptodactyly | No | 6/8 (75%) |
| Pregnancy complications | Polyhydramnios | Intrauterine growth restriction | Microcephaly, ncreased nuchal thickness, flattened forehead | Oligohydramnios | No | Intraventricular hemorrhage, Intrauterine demise | Perinatal stroke | IVF conception with vanishing twin at 12-16 weeks | N/A | N/A | 7/8 (87.5%) |

